# Relationship between common polymorphisms of NRAMP1 gene and pulmonary tuberculosis in Lorestan LUR population

**DOI:** 10.1101/2022.05.01.22274504

**Authors:** Ali Amiri, Toomaj Sabooteh, Farhad Shahsavar

**Author notes:** **Corresponding author:** Farhad Shahsavar, Hepatitis Research Center, Lorestan University of Medical Sciences, Khorramabad, Iran.

## Abstract

**Introduction:** Tuberculosis (TB) is caused by Mycobacterium tuberculosis. In humans, a number of genes have been identified as susceptible to pulmonary tuberculosis. The relationship between NRAMP1 polymorphisms and pulmonary tuberculosis has been studied in different populations and has reported contradictory results. The aim of this study was to investigate the relationship between the common polymorphisms of NRAMP1 gene and the susceptibility to pulmonary tuberculosis in the LUR Population of Lorestan province of Iran.

**Materials and Methods:** In this case control study, three common polymorphisms of NRAMP1 gene (3’UTR, INT4 and D543N) were genotyped using PCR-RFLP technique in the LUR population of Lorestan province. In this study, 100 patients with pulmonary tuberculosis (PTB) were studied as case group and 100 healthy controls that matched for age and sex with the patient group, studied as control group. Statistical analysis was performed using SPSS 18 software.

**Results:** In the present study we observed that the GG genotype of D543N polymorphism was statistically significantly associated with increased susceptibility to TB (84% in the case group vs. 72% in the control group, %95CI=1.024-4.071, OR=2.042, P=0.0405). Also, G allele of D543N polymorphism was statistically significantly associated with increased susceptibility to pulmonary tuberculosis (90% in the case group vs. 81.5% in the control group, %95CI=1.140-3.663, OR=2.043, P=0.015). On the other hand, the frequency of allele A of D543N polymorphism was significantly lower in patients than in the control group (10% in the case group vs. 18.5% in the control group, %95CI=0.273-0.878, OR=0.489, P=0.015). Although genotypic and allelic frequency of 3’UTR and INT4 polymorphisms between patients and controls showed no significant differences in the study population.

**Discussion and conclusion:** Our observations showed that GG genotype and G allele of D543N polymorphism have a significant role in increasing the susceptibility to pulmonary tuberculosis in the LUR Population of Lorestan province. Also, allele A of D543N polymorphism has a significant effect on resistance to pulmonary tuberculosis in this population. Although there was no significant correlation between genotypes and alleles of 3’UTR and INT4 polymorphisms with susceptibility to or resistance to pulmonary TB in this population. It is suggested that a larger sample size be used in future studies. It is also recommended to conduct this type of study on other ethnicities.

## Introduction

Pulmonary tuberculosis (TB) is a chronic infectious disease that is one of the most important causes of death in the developing countries. TB is caused by Mycobacterium tuberculosis and it affects the lungs. The WHO has reported that 1.5 million people have died from pulmonary tuberculosis in 2020. Pulmonary tuberculosis, with 8.8 million new infections each year, after COVID-19 and even more than HIV/AIDS, is the second leading infectious cause of death and the 13th leading cause of death in the world (1). About one-third of the world’s people have been infected with Mycobacterium tuberculosis, but only 5-10% of them will develop active pulmonary tuberculosis (2). According to the WHO report, 5.6 million men, 3.3 million women and 1.1 million children have been diagnosed with pulmonary tuberculosis in 2020. Although pulmonary tuberculosis has been observed in all age groups and in all countries of the world, on the other hand, it is preventable and curable (3). In various studies, it has been reported that most people who have contracted tuberculosis reside in China, Pakistan, South Africa, India, the Philippines and Nigeria (4). This suggests that genetic and ethnic differences may play a role in susceptibility to pulmonary tuberculosis. The most important risk factors for pulmonary tuberculosis include alcohol abuse, diabetes, HIV, chronic corticosteroid treatment, malnutrition, advanced age, weakened immunity, socioeconomic status, and genetic factors (5).

The role of genetic factors in susceptibility to pulmonary tuberculosis is still inconclusive. Exposure to these risk factors was not a definite reason for pulmonary tuberculosis, therefore, genetic factors such as single nucleotide polymorphisms may play an effective role in pulmonary tuberculosis. Also, some studies have shown that genetic susceptibility of individuals combined with environmental factors may play a role in the mechanism of pulmonary tuberculosis infection (6).

In several studies, the role of different genes in susceptibility to pulmonary tuberculosis has been investigated, indicating the role of ethnicity in the disease. It has also been reported that these genetic factors are associated with the severity of the disease and concurrent infection with various diseases (especially HIV). Most of these genes are localized in the loci involved in the immune response, which shows that people’s genetic background is effective in the immune response and the potential for pulmonary tuberculosis (7). One of which is KIR3DS1 gene and it combines with HLA-B Bw4 and Ile80 ligand (8,9). Also, other studies showed Toll like Receptors (TLRs) (10-14), Mannose Binding Lectin (MBL) (15), Purinergic receptor P2X, ligand-gated ion channel 7 (P2X7) (16), Interlukin-10 (IL-10) (17) and Tumor Necrosis Factor-α (TNF-α) (18), vitamin D receptor (VDR) (19) has roles in the susceptibility or resistance to TB. When a person is infected with Mycobacterium tuberculosis, the patient’s immune system attempts to eliminate the pathogen by producing defensive protein molecules and recruiting phagocytic cells.

Natural resistance-associated macrophage protein 1 (NRAMP1) is also referred to as the solute carrier family 11 proton-coupled divalent metal ion transporter membrane1 (SLC11A1), which it plays an important role in the creation of an immune response in pulmonary tuberculosis (13,20). NRAMP1 encodes a divalent transition metal (Fe and Mn) transporter that is located on the lysosomal membrane (21). In addition Iron has a crucial role in producing reactive oxygen and nitrogen intermediates in macrophages and is an important mycobacterial nutrient (22).

Various studies have shown that NRAMP1 is associated with various inflammatory and infectious diseases. They include Leishmania, Salmonella, and Mycobacteria. Immune cytochemical studies show that NRAMP1 protein exhibits pleiotropic effects on macrophage activation, including up-regulation of MHC II molecules, increased production of ROS, increased expression of iNOS (inducible NO synthase), reactive nitrogen intermediates involved in the oxidative burst, increased production of the proinflammatory cytokines, such as IL1-β and TNFα, regulation of chemokines and increased NO release (23), regulation of intracellular membrane vesicle trafficking of (24,25), regulation of cytoplasmic cation levels that promotes the generation of toxic hydroxyl radicals, which significantly contribute to the killing of intracellular pathogens (26), Although it is not yet known exactly the function of human NRAMP1 protein.

NRMAP1 polymorphisms contains rs17235416 (3’UTR), rs17235409 (D543N), rs3731865 (INT4), and rs34448891 (5=(GT)n). In several studies, the relationship between NRAMP1 polymorphisms and risk of pulmonary tuberculosis (PTB) has been investigated, however, due to low sample size in some studies and different ethnicities, the results of these studies are inconsistent and inconclusive (27-52). In a study conducted by Medapati et al (52) in the Indian population, the results showed a significant relationship between NRAMP1 3’UTR polymorphism and susceptibility to pulmonary tuberculosis. However, no significant relationship was observed in the study of Jafari et al (51).

In studies conducted in West African populations, there was a significant relationship between NRAMP1 gene polymorphisms (3’UTR, INT4 and D543N) and susceptibility to pulmonary tuberculosis in some populations including Gambian population (53). While there was no significant relationship between Some populations including Thai (34), Brazilians (54), Taiwanese (28) and Moroccans (55).

The present study aimed to investigate 3 polymorphisms in NRAMP gene: INT4 (469+14G>C) rs3731865, D543N (codon 543 Asp to Asn) rs17235409 and 3’UTR (1729 + 55del4) rs17235416, in relation to the susceptibility to TB in LUR population residents of Lorestan Province of Iran. Identification of host genes in different ethnicities can be effective in identifying the role of ethnic differences in susceptibility to some infectious diseases. It may also play an effective role in our knowledge of pathogenesis, prevention and treatment of pulmonary tuberculosis. This article is written, based on the STROBE statement guidelines for reporting observational studies (supplementary file 1) (56).

## Materials and Methods

### Study design

In this case control study, the number of patients in the case group and the control group were determined using the statistical formula and based on previous studies, 100 subjects were assigned to each group. The statistical population of the patient group included all patients with pulmonary TB who referred to Khorramabad Clinic of Health, whose disease was confirmed by histological and sputum culture. case group, was 100 Iranian unrelated LUR patients with Pulmonary Tuberculosis were referred to Khorramabad Health Center. All patients received standard tuberculosis treatment, and none had drug resistance.

The statistical population of the control group was selected from healthy individuals of the LUR population in the Lorestan province. The control group was included in the study by simple sampling (accessible) and based on age and gender matched with patient population. The control group had no history of pulmonary tuberculosis and no evidence of previous radiographic changes of the chest associated with tuberculosis. All subjects under study had parents of the same race. Individuals who had any of the exclusion criteria were excluded from the study.

### Inclusion and exclusion criteria

The Inclusion criteria were: patients with newly diagnosed pulmonary tuberculosis, having lur ethnicity, having pulmonary tuberculosis symptoms, Sputum smear-positive, A chest radiograph of active disease, satisfaction to participate in the study. Also, people with diabetes mellitus, ischemic heart disease, chronic renal failure, any chronic inflammatory disease, jaundice, hepatitis C, HIV, any autoimmune disease, immunosuppressive drugs, HBsAg positive, and People who did not agree to participate in the study were excluded.

### DNA Extraction

After obtaining informed written consent, from 100 patients with pulmonary tuberculosis and 100 healthy controls who entered the study, 2 ml of peripheral blood was taken in the tubes containing EDTA anticoagulant. After transferring the samples to the Laboratory of Immunogenetics, using the kit QIAmp (Qiagen-Germany) according to the manufacturer’s protocol, DNA was extracted genomic samples of peripheral blood leukocytes. After quantitative and qualitative measurements, samples of DNA, to the time of testing were stored at −70 ° C.

### Genotyping by PCR-RFLP

In this study, the most common polymorphisms of the NRAMP1 gene, INT4 (469 + 14G/C) (rs3731865), 3’UTR (1729 + 55del4) (rs17235416) and D543N (codon 543, Asp to Asn) (rs17235409) Were examined in the patients and control groups. The temperature conditions of the PCR reaction are listed in Table 1. PCR amplification was performed using pure Mastercycler DNA (BioRad, USA) in a 20μl reaction volume.

**Table 1.** Thermal conditions of PCR reaction (57)

| Number of cycles | Steps | Duration | Temperature |
| --- | --- | --- | --- |
| 1 | Initial denaturation | 5 minutes | 94 °C |
| 35 | denaturation | 30 seconds | 95 °C |
|  | Annealing | 30 seconds | 52 °C |
|  | Extension | 30 seconds | 72 °C |
| 1 | Final elongation | 10 minutes | 72 °C |

To determine the common polymorphisms of the NRAMP1 gene, used, the PCR-RFLP technique previously used by Medapati et al. In 2017 (57), Nugraha et al. In 2011 (58), Hanta et al. In 2012 (59) and Farnia et al. In 2008 (38). Forward and Reverse primer sequences NRAMP1 gene polymorphisms that were designed in previous studies, were used. These sequences are listed in Table 2.

**Table 2:**
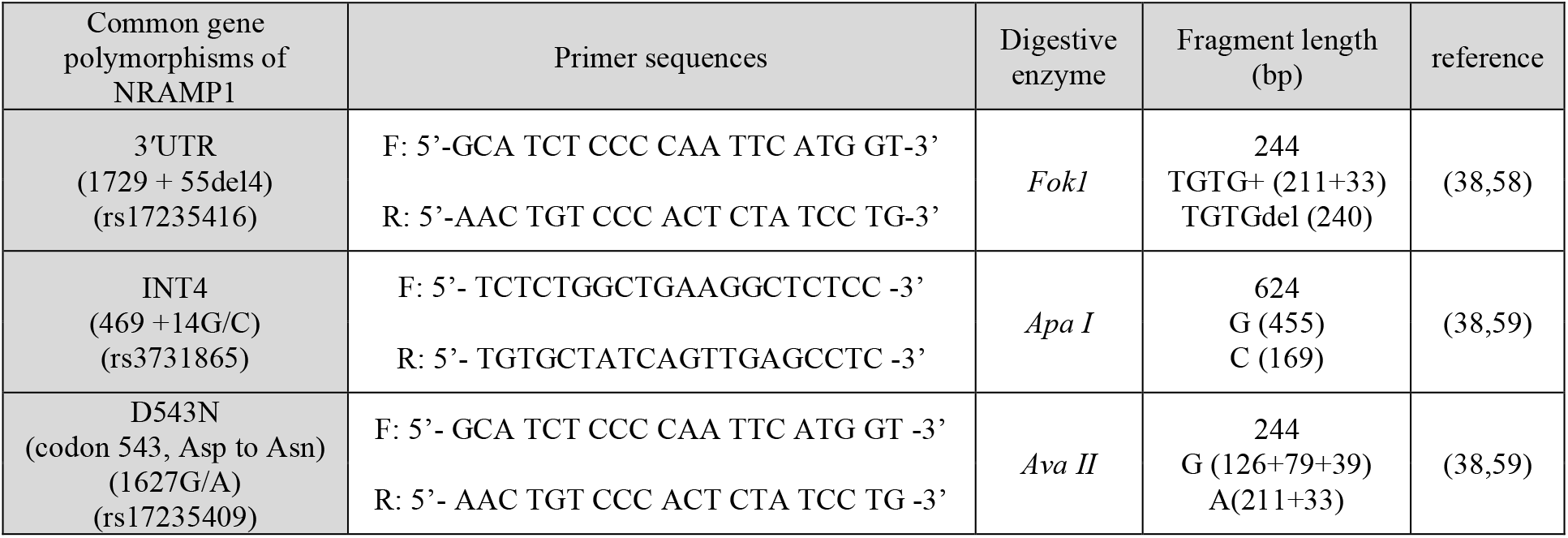
Primer sequences of NRAMP1 gene polymorphisms

In order to perform RFLP for 3’UTR polymorphism, 20 μl of amplicon was digested with 5 units of Fok1 digesting enzyme at 37°C for 2 hours. For the TGTG + allele, the length of the fragments was 33+211 bp, and for the TGTGdel allele, the length of the fragments was 240 bp (38,57).

In order to perform RFLP for INT4 polymorphism, 20 μl of amplicon was digested with 5 units of Apa I digesting enzyme at 37°C for 2 hours. For the G allele, the length of the fragments was 455 bp, and for the C allele, the length of the fragments was 169 bp (38,59).

In order to perform RFLP for D543N polymorphism, 20 μl of amplicon was digested with 5 units of Ava II digesting enzyme at 37°C for 2 hours. For the G allele, the length of the fragments was 126+79+39 bp, and for the A allele, the length of the fragments was 211+33 bp (38,59).

The proliferated PCR product was electrophoresed on 2% agarose gel containing ethidium bromide. Bands, under ultraviolet light, appeared. Genotypes were determined according to the pattern of the produced part. Finally, to ensure the correct result of the PCR, we sent 5% of the samples randomly to determine the DNA sequence.

### Statistical Analysis

After data collection, the data were entered into SPSS 18 software and statistical analysis was performed. Genotypic and allelic frequencies of common polymorphisms of NRAMP1 gene (INT4 (469 + 14G/C) (rs3731865), 3’UTR (1729 + 55del4) (rs17235416) and D543N (codon 543, Asp to Asn) (rs17235409) in the patient group and control group was determined by direct counting. Departure from Hardy-Weinberg equilibrium and frequency of all polymorphisms in the patient group and the control group were determined. Using descriptive statistics (frequency and percentages), the results were presented in statistical tables and Analytical statistics (t-test and chi-square) were used to measure the association and the effect of variables. Differences in genotypic and allelic distribution of NRAMP1 gene polymorphisms between patient group and control group were calculated by Chi-Square test and Fisher’s exact test. Finally, P<0.05 was considered statistically significant. Also, in the analysis of information, odds ratio (OR) and 95% confidence interval (CI) were used.

### Ethical considerations

The protocol of this study was approved by the Ethics Committee in the research of Lorestan University of Medical Sciences with the code of ethics IR.LUMS.REC.1395.114. Entry into the study was voluntary. Before the beginning of the study, informed consent was obtained from each patient for the use of patient information in relevant research. Also, all patient information was considered as a secret, and any disclosure was refused. Identification codes were used to prevent the registration of patients’ names and last names. The data were confidential and recorded as SPSS software by numerical codes.

### Limitations

The limitations and problems of this study can be pointed to problems of sampling from healthy individuals.

## Results

In this case-control study, that disigned to investigate the association of common polymorphisms of NRAMP1 gene with pulmonary tuberculosis in Lur population in Lorestan province of Iran, 100 patients with smear positive pulmonary tuberculosis were evaluated as the case group and 100 healthy subjects as control group were studied. The case group consisted of 46 males and 54 females, and the control group consisted of 52 males and 48 females. The mean age of the case group was 34.73 ± 3.21(in the age range of 26 to 57 years old) and the mean age of the control group was 31.12 ± 2.25 (in the age range of 23 to 61 years old). Genotypic and allelic distribution of NRAMP1 polymorphisms were not diverted from Hardy-Weinberg equilibrium in all samples (P>0.05). The genotypic frequency of 3’UTR, INT4 and D543N polymorphisms, and the association of each with pulmonary tuberculosis susceptibility is presented in Table 3.

**Table 3.**
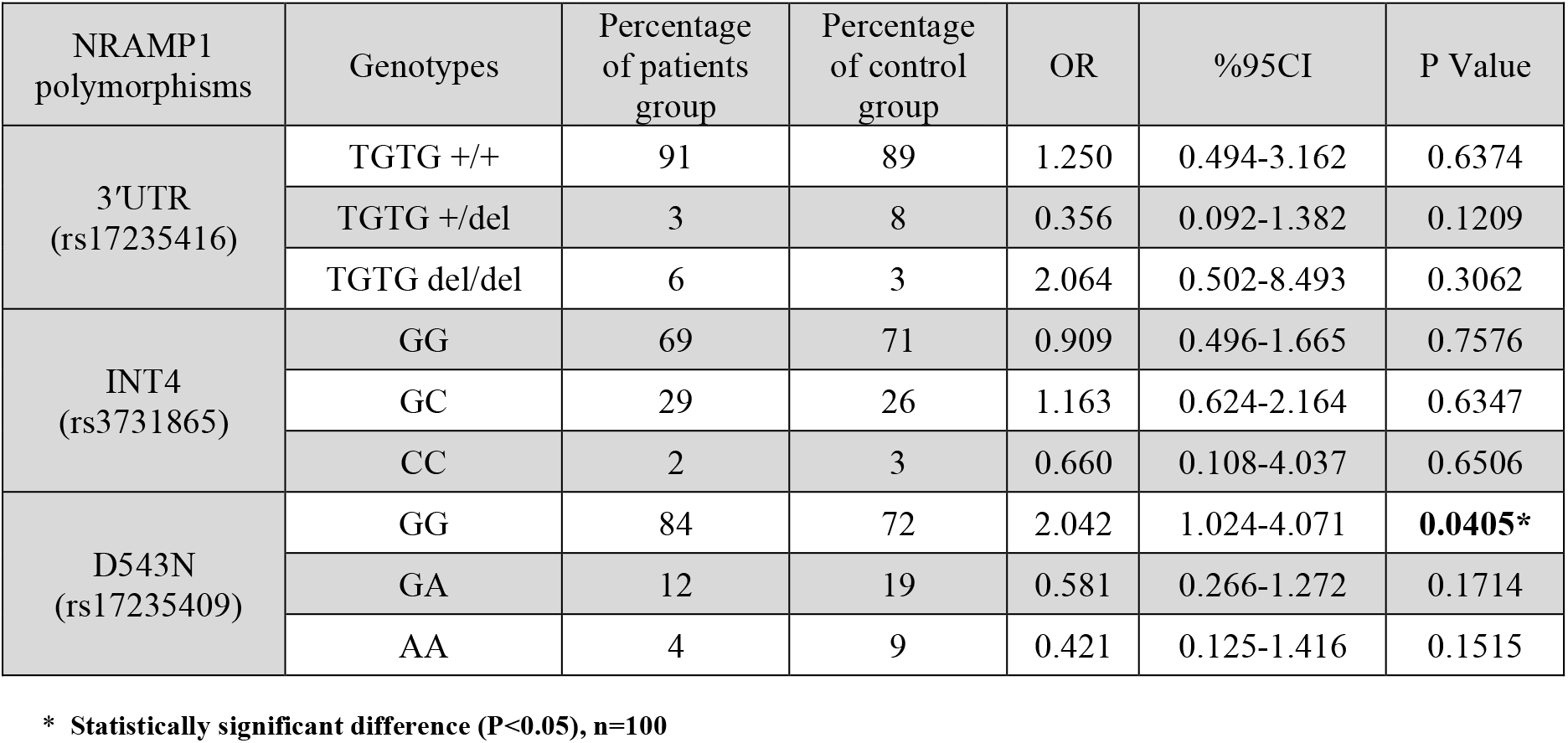
Genotype distribution of NRAMP1 polymorphisms in patients and controls

As shown in Table 3, for the 3’UTR polymorphism, the frequency of TGTG +/+, TGTG +/del and TGTG del/del genotypes in patient group was 91%, 3% and 6%, respectively, and in the control group was 89%, 8% and 3%, respectively. for the INT4 polymorphism, the frequency of GG, GC and CC genotypes in patient group was 69%, 29% and 2%, respectively, and in the control group was 71%, 26% and 3%, respectively. for the D543N polymorphism, the frequency of GG, GA and AA genotypes in patient group was 84%, 12% and 4%, respectively, and in the control group was 72%, 19% and 9%, respectively.

GG genotype of D543N polymorphism was statistically significantly associated with increased susceptibility to pulmonary tuberculosis (84% in the case group vs. 72% in the control group, 95%CI= 1.024-4.071, OR=2.042, P=0.0405). This genotype almost doubles the chance of having pulmonary tuberculosis in the population under study (OR=2.042). The genotypic frequency of 3’UTR and INT4 polymorphisms was not significantly different between the patients and the control group.

The allelic frequency of 3’UTR, INT4 and D543N polymorphisms, and the association of each with pulmonary tuberculosis susceptibility is presented in Table 4.

**Table 4.** Allelic distribution of NRAMP1 polymorphisms in patients and controls

| NRAMP1 polymorphisms | Alleles | Percentage of patients group | Percentage of control group | OR | %95CI | P Value |
| --- | --- | --- | --- | --- | --- | --- |
| 3'UTR (rs17235416) | TGTG + | 92.5 | 93 | 0.928 | 0.436-1.978 | 0.8471 |
|  | TGTG del | 7.5 | 7 | 1.077 | 0.506-2.295 | 0.8471 |
| INT4 (rs3731865) | G | 83.5 | 84 | 0.964 | 0.567-1.640 | 0.8922 |
|  | C | 16.5 | 16 | 1.037 | 0.610-1.765 | 0.8922 |
| D543N (rs17235409) | G | 90 | 81.5 | 2.043 | 1.140-3.663 | **0.015\*** |
|  | A | 10 | 18.5 | 0.489 | 0.273-0.878 | **0.015\*** |

\* Statistically significant difference (P<0.05), n=100

As shown in Table 4, for the 3’UTR polymorphism, the frequency of TGTG + and TGTG del alleles in patient group was 92.5% and 7.5%, respectively, and in the control group was 93% and 7%, respectively. for the INT4 polymorphism, the frequency of G and C alleles in patient group was 83.5% and 16.5%, respectively, and in the control group was 84% and 16%, respectively. for the D543N polymorphism, the frequency of G and A alleles in patient group was 90% and 10%, respectively, and in the control group was 81.5% and 18.5%, respectively.

G allele of D543N polymorphism was statistically significantly associated with increased susceptibility to pulmonary tuberculosis (90% in the case group vs. 81.5% in the control group, 95%CI= 1.140-3.663, OR=2.043, P=0.015). This allele almost doubles the chance of having pulmonary tuberculosis in the population under study (OR=2.043).

The frequency of allele A of D543N polymorphism was significantly lower in patients than control group (10% in the case group vs. 18.5% in the control group, 95%CI= 0.273-0.878, OR=0.489, P=0.015). This allele 51% reduces the chance of having pulmonary tuberculosis in the population under study (OR=0.489).

The allelic frequency of 3’UTR and INT4 polymorphisms was not significantly different between the patients and the control group.

## Discussion

Pulmonary tuberculosis is one of the common infectious diseases that annually causes more than 3 million deaths worldwide. The causative agent of this disease is Mycobacterium tuberculosis. Despite the implementation of extensive control programs over the past two decades, pulmonary tuberculosis remains a high-risk infectious disease. On the other hand, the emergence and expansion of drug-resistant strains has created new concerns globally. Studies have shown that the sensitivity to the disease, and even its course and its trends among different people, is different and any person exposed to this bacterium does not necessarily have tuberculosis. These differences can be due to host factors, especially the genetic susceptibility of different individuals to the disease (60).

So far, no study has been done to investigate the role of common NRAMP1 gene polymorphisms with susceptibility to pulmonary tuberculosis in the Lur population of Lorestan province. This study was developed and implemented in the Lur population of Lorestan province with the aim of investigate the role of common NRAMP1 gene polymorphisms (INT4 (469 + 14G/C) (rs3731865), 3’UTR (1729 + 55del4) (rs17235416) and D543N (codon 543, Asp to Asn) (rs17235409) in susceptibility to pulmonary tuberculosis.

In the present study, we observed that GG genotype of D543N polymorphism was statistically significantly associated with increased susceptibility to pulmonary tuberculosis. This genotype almost doubles the chance of having pulmonary tuberculosis in the population under study. G allele of D543N polymorphism was statistically significantly associated with increased susceptibility to pulmonary tuberculosis. This allele almost doubles the chance of having pulmonary tuberculosis in the population under study. The frequency of allele A of D543N polymorphism was significantly lower in patients than control group. This allele 51% reduces the chance of having pulmonary tuberculosis in the population under study. The genotypic and allelic frequency of 3’UTR and INT4 polymorphisms was not significantly different between the patients and the control group.

Various studies have been done to investigate the role of common NRAMP1 gene polymorphisms with susceptibility or resistance to pulmonary tuberculosis in different populations, which reported different results.

### 3′UTR Polymorphism

One of the objectives of the present study was to investigate the relationship between genotypes and alleles of 3’UTR polymorphism in susceptibility to or resistance to pulmonary tuberculosis in the studied population. As it was observed, there was not statistically significant relationship between any of the genotypes and alleles of TGTG +/+ (P=0.63), TGTG +/del (P=0.12), TGTG del/del (P=0.30), TGTG + (P=0.84) and TGTG del (P=0.84) with susceptibility to or resistance to pulmonary tuberculosis in the studied population. These results were consistent with the results of some similar studies. For example, in 2010, a study by Hatta et al. Was conducted on 58 patients with pulmonary tuberculosis and 198 healthy individuals in Indonesia using PCR-RFLP techniques. In their study, there was no significant correlation between genotypes and alleles of 3’UTR polymorphism with susceptibility to or resistance to pulmonary tuberculosis (%95CI=0.484-1.693, OR=0.905, P=0.755) (41). Also, in the study of Stagas et al. In 2011, no significant correlation was found between genotypes and alleles of 3’UTR polymorphism with susceptibility to pulmonary tuberculosis (61). In 2016, a case-control study was performed by Jafari et al. On 94 patients with pulmonary TB and 122 healthy individuals from Iranian population using ARMS-PCR technique. In their study, there was no statistically significant correlation between genotypes (%95CI=0.22-2.13, OR=0.69, P=0.711) and alleles (%95CI=0.23-2.12, OR=0.70, P=0.718) of 3’UTR polymorphism with susceptibility to pulmonary tuberculosis (51). Also in 2015, a study by Trifunovic et al. Was conducted on 110 patients with pulmonary TB and 67 healthy people from the Serbian population using PCR-RFLP technique. They observed in their study that there was no significant correlation between genotypes and alleles of 3’UTR polymorphism with susceptibility to pulmonary disease (%95CI=0.012-31.061, OR=0.609, P=0.99) (62).

On the other hand, some studies have reported a significant correlation between 3’UTR polymorphism and pulmonary tuberculosis susceptibility. For example, a study by Ghozali et al in 2016 was conducted on 40 patients with pulmonary TB and 50 healthy individuals with PCR-RFLP technique. In their study, they reported a significant correlation between the TGTG del allele of 3’UTR polymorphism and the susceptibility to pulmonary tuberculosis (78.8% in the patient group vs. 51% in the control group, %95CI=1.83-6.9, OR=3.56, P=0.002) (63). Also, in 2017, a case-control study was conducted by Medapati et al. In the Indian population using the PCR-RFLP technique. In their study, there was a statistically significant correlation between genotypes (%95CI=1.01-8.81, OR=2.99, P=0.005) and alleles (%95CI=2.03-14.57, OR=5.44, P=0.001) of 3’UTR polymorphism with increasing susceptibility to pulmonary TB in the Indian population (52).

In 2011, a study by Nugraha et al. Was conducted on 69 patients with pulmonary TB and 43 healthy individuals from the Indonesian population. They observed in their study that there was a significant correlation between TGTG del/del genotype 3’UTR polymorphism and the susceptibility to pulmonary tuberculosis (29% in the patient group vs. 5% in the control group, P=0.0072) (44). The results of meta-analysis study that was conducted in 2012 by Meilang and colleagues found that, in general, in all races studied 3’UTR polymorphism was significantly associated with increased susceptibility to pulmonary tuberculosis (%95CI=1.25-1.68, OR=1.45, P<0.05) (64).

### INT4 Polymorphism

Another aim of the present study was to investigate the relationship between genotypes and alleles of INT4 polymorphism in susceptibility to or resistance to pulmonary tuberculosis in the studied population. As it was observed, there was not statistically significant relationship between any of the genotypes and alleles of GG (P=0.75), GC (P=0.63), CC (P=0.65), G (P=0.89) and C (P=0.89) with susceptibility to or resistance to pulmonary tuberculosis in the studied population. These results were consistent with the results of some similar studies. For example, in the study of Nugraha et al. In 2011, there was no statistically significant relationship between any of the genotypes and alleles of INT4 polymorphism with susceptibility to pulmonary tuberculosis (P=0.370) (44). It was also reported in the Trifunovic et al. Study in 2015 that there is no statistically significant correlation between genotypes and alleles of INT4 polymorphism in susceptibility to pulmonary tuberculosis (%95CI=0.809-2.878, OR=1.526, P=0.21) (62). In the study of Jafari et al. In 2016, no significant association was found between genotypes (%95CI=0.69-2.21, OR=1.23, P=0.576) and alleles (%95CI=0.70-2.02, OR=1.19, P=0.610) of INT4 polymorphism and susceptibility to pulmonary tuberculosis in Iranian population (51). Also, in the study of Hatta et al in 2010, there was no significant correlation between genotypes and INT4 polymorphism alleles with susceptibility to pulmonary tuberculosis (%95CI=0.373-2.536, OR=0.973, P=0.955) (41).

On the other hand, some studies have reported a significant association between INT4 polymorphism and pulmonary tuberculosis. For example, in a meta-analysis study conducted by Meilang et al. In 2012, it was reported that in general, the INT4 polymorphism was significantly associated with increasing the susceptibility to pulmonary tuberculosis (%95CI=1.09-1.49, OR=1.27, P<0.05) (64). Also, in the study of Ghozali et al in 2016, was observed significant correlation between C allele of INT4 polymorphism and pulmonary tuberculosis (10% in the patient group vs. 2% in the control group, %95CI=1.1-26.4, OR=5.44, P=0.02) (63). In 2011, another study by Stagas et al. Was conducted on 142 patients with pulmonary TB and 144 healthy people from the Egyptian population. In their study, they observed that the CC genotype of INT4 polymorphism had a statistically significant correlation with the increased risk of pulmonary tuberculosis in the Egyptian population (61). Also, in 2017, a systematic review and meta-analysis was conducted by Yuan et al. They reported that in general and in the comprehensive analysis of studies conducted in different races, INT4 polymorphism significantly increased the susceptibility to pulmonary tuberculosis (%95CI=1.04-1.75, OR=1.35, P=0.02). It was also observed that in the Africans, there is a significant correlation between GC genotype (%95CI=1.14-2.04, OR=1.53, P=0.004) and C allele (%95CI=1.08-1.85, OR=1.41, P=0.012) of INT4 polymorphism, with increasing susceptibility to pulmonary tuberculosis (65).

### D543N Polymorphism

Another aim of the present study was to investigate the relationship between genotypes and alleles of D543N polymorphism in susceptibility to or resistance to pulmonary tuberculosis in the studied population. As it was observed, there was a significant correlation between GG genotype (P=0.04) and G allele (P=0.01) with increasing susceptibility to pulmonary tuberculosis. Also, there was a significant association between allele A (P=0.01) and resistance to pulmonary tuberculosis in the studied population. Although there was no significant relationship between GA (P=0.63) and AA (P=0.65) genotypes with susceptibility to or resistance to pulmonary TB in the studied population. These results were consistent with the results of some similar studies. For example, in a meta-analysis study by Meilang et al. In 2012, it was reported that in general, in all of the studied races, D543N polymorphism has a significant correlation with the increase susceptibility to pulmonary tuberculosis (%95CI=1.11-1.55, OR=1.31, P<0.05) (64).

On the other hand, it has been reported in some studies that there is no significant relationship between D543N polymorphism and pulmonary tuberculosis. For example, in the study of Ghozali et al in 2016, no significant association was found between allele A of D543N polymorphism and susceptibility to pulmonary tuberculosis (16.3% in the patient group vs. 15% in the control group, P=0.84). Also, in their study, there was no significant correlation between GA genotype of D543N polymorphism and susceptibility to pulmonary tuberculosis (32.5% in the patient group vs. 30% in the control group, P=0.79) (63). Also, in the study of Stagas et al. In 2011, no significant correlation was found between genotypes and alleles of D543N polymorphism with susceptibility to pulmonary tuberculosis (P=0.97) (61). In a systematic and meta-analysis study by Yuan et al in 2017, there was no significant relationship between D543N polymorphism and susceptibility to Pulmonary tuberculosis in general and in a comprehensive analysis of studies in different races. Although in the subgroup analysis based on race, there was a significant correlation between GA genotype (%95CI=1.03-1.67, OR=1.31, P=0.03) and allele A (%95CI=1.02-1.63, OR=1.29, P=0.032) of D543N polymorphism in American race, with an increased risk of pulmonary tuberculosis (65). Also, in the study of Jafari et al. In 2016, no significant association was found between genotypes (%95CI=0.20-1.86, OR=0.62, P=0.551) and alleles (%95CI=0.21-1.86, OR=0.63, P=0.395) of D543N polymorphism and susceptibility to pulmonary tuberculosis (51). In the study of Trifunovic et al. In 2015, there was no statistically significant relationship between genotypes and alleles of D543N polymorphism in susceptibility to pulmonary disease (%95CI=0.041-37.14, OR=1.229, P=0.61) (62). In a study by Hatta et al in 2010, there was no significant correlation between genotypes and alleles of D543N polymorphism with susceptibility or resistance to pulmonary tuberculosis (%95CI=0.833-10.119, OR=2.904, P=0.082) (41). Also, in the study of Nugraha et al. In 2011, there was no statistically significant relationship between any genotypes and alleles of D543N polymorphism with susceptibility to pulmonary tuberculosis (P=0.098) (44).

## Conclusion

In the present study it was observed that GG genotype and allele G of the D543N polymorphism of NRAMP1 gene, significantly increased susceptibility to pulmonary tuberculosis in Lur population residents in lorestan province of iran. Also, allele A of D543N polymorphism of NRAMP1 gene is also effective in resistance to pulmonary tuberculosis in this population. No significant correlation was found between the genotypes and alleles of the 3’UTR and INT4 polymorphisms of the NRAMP1 gene and the susceptibility to or resistance to pulmonary tuberculosis in this population. These results were in line with the results of some studies that were carried out for this purpose on other populations, while not consistent with the results of some other studies. The contradictory results reported by various studies are not unpredictable because various factors such as variation in sample size, differences in the intensity of the relationship in different studies, different genotyping techniques, also different races, can lead to differences in results.

It is suggested that in future studies, this type of study should be done with a larger sample size and with different genotyping techniques, on the Lur and other Iranian ethnicities. It is also suggested that similar studies be conducted to investigate the association of polymorphisms of other genes involved in susceptibility or resistance to pulmonary tuberculosis, that So far, their role in susceptibility or resistance to tuberculosis in different ethnic groups had conflicting results.

## Data Availability

All data produced in the present study are available upon reasonable request to the authors.

## Acknowledgments

This article is a part of Dr. Sanaz Rostami’s thesis on General Medicine, that research project was approved by the Research Council of Medicine School at Lorestan University of Medical Sciences, Grant number: 134. This research was conducted with financial assistance from the Deputy of Research and Technology of Lorestan University of Medical Sciences. We would like to thank all those who helped us in this way, especially Dr. Khatereh Anbari, because of statistical counseling of this research project. Other parts of this thesis will be published in our next articles and according to COPE guideline, the authors’ names will be different in the next articles.

## Contributors

The following individuals have contributed: **Concept and Design:** Ali Amiri, Farhad Shahsavar, Toomaj Sabooteh; **Data Acquisition:** Ali Amiri, Farhad Shahsavar, Toomaj Sabooteh, Sanaz Rostami; **Data Analysis:** Khatereh Anbari, Toomaj Sabooteh; **Drafting the Manuscript:** Toomaj Sabooteh; **Critical Revising of the Manuscript:** Ali Amiri, Farhad Shahsavar; **Final Approval of the Manuscript:** All Authors.

